# Protective targets of PfSPZ vaccines identified from whole-genome sieve analysis of isolates from malaria vaccine efficacy trials in West Africa

**DOI:** 10.1101/2025.03.04.25323352

**Authors:** Ankit Dwivedi, Ryan J. Scalsky, David G. Harris, Thomas C. Stabler, Biraj Shrestha, Sudhaunshu Joshi, Chakshu Gandhi, James B. Munro, Olukemi O. Ifeonu, Alphonse Ouedraogo, Alfred B. Tiono, Drissa Coulibaly, Amed Ouattara, Thomas L. Richie, B. Kim Lee Sim, Christopher V. Plowe, Kirsten E. Lyke, Shannon Takala-Harrison, Stephen L. Hoffman, Mahamadou A. Thera, Sodiomon B. Sirima, Matthew B. Laurens, Joana C. Silva

## Abstract

Identification of antigens targeted by a protective response is a central quest in malaria vaccinology. Whole-genome sieve analysis (SA_WG_) in samples collected from placebo-controlled field trials of *Plasmodium falciparum* (Pf) sporozoite (SPZ) vaccines may enable identification of Pf pre-erythrocytic antigens. We applied SA_WG_ to genomic data generated from Pf isolates collected during two field trials measuring the efficacy, in malaria-exposed African adults, of two PfSPZ vaccines. These randomized, double-blind, placebo-controlled trials were conducted in regions of Mali and Burkina Faso characterized by high seasonal transmission, where parasite genetic diversity is high. Genomic sites in which the vaccine allelic state was significantly underrepresented among breakthrough infections in vaccinees relative to placebo recipients were termed “target sites”. Protein-coding loci containing target sites that changed amino acids were termed “target loci”. The SA_WG_ conducted on clinical trial samples from the Burkina Faso and Mali trials identified 138 and 80 single-copy protein-coding target loci in the Burkinabe and Malian data sets, respectively, with twelve common to both, a number significantly higher than expected (*E* = 3.9; 99%CI = [0, 9]). Among these was the thrombospondin-related anonymous protein locus, which encodes PfSSP2|TRAP, one of the most abundant and well-characterized pre-erythrocytic stage antigen as well as other genes encoding membrane-associated proteins of unknown function. These results identify SA_WG_ as a potentially powerful tool for identifying protective vaccine antigens in recombining pathogens with large genome size and reveals potential new protective Pf antigens.

## INTRODUCTION

The World Health Organization (WHO) recommended the very first malaria vaccine, RTS,S|AS01, in October 2021, reflecting decades of research. The formulation of RTS,S|AS01 is based on the variant of *Plasmodium falciparum* (Pf) circumsporozoite protein (CSP) encoded by the 3D7 strain. In October 2023, a second malaria vaccine, R21|Matrix-M, based on the same variant of CSP, was endorsed by the WHO.^1^ While this momentous advance will reduce malaria morbidity and mortality, it is clear that neither vaccine will meet the current WHO strategic goal of achieving >90% efficacy against Pf infection.^2^ Furthermore, RTS,S|AS01 has been shown to induce allele-specific protection^3^ and is, therefore, susceptible to vaccine escape, highlighting the need for additional vaccines to protect against breakthrough infections.

Whole organism vaccines based on Pf sporozoites (PfSPZ) are in late stage development,^4^ and are highly promising as pre-erythrocytic candidate vaccines that can prevent malaria disease and interrupt malaria transmission by preventing parasites from ever leaving the liver. They possess highly desirable features,^5^ including (i) the stimulation of human immunity to multiple Pf antigens present during the pre-erythrocytic phase of parasite development; ii) up to 100% protection against controlled human malaria infection (CHMI) with parasites homologous^6,7^ and heterologous^8^ to the vaccine parasite strain; and (iii) durable efficacy without boosting in malaria-experienced adults in highly endemic settings in sub-Saharan Africa.^9–11^ These features represent an extremely high achievement for a malaria vaccine that must overcome a lifetime of immune exhaustion resulting from repeat malaria infection^12–14^ to stimulate a protective antimalarial human immune response. Efficacy in adults living in a high transmission setting with limited ability to respond strongly to a malaria vaccine suggests that PfSPZ vaccines may achieve higher efficacy in pediatric populations with less preexisting immunity to malaria. Nevertheless, the overall lower protection against heterologous relative to homologous CHMI with the same dose of PfSPZ^15^ is also consistent with genotype- (or allele)- specific efficacy.

Knowledge of Pf antigens that induce a protective response is required to advance malaria subunit vaccine development and could be used to enhance PfSPZ vaccines. New multivalent subunit vaccines or multi-strain PfSPZ vaccines, where the multiple components complement each other, may reduce or eliminate vaccine escape and provide broad, strain-transcending protection against malaria. As we recently outlined, allele-specific efficacy, while detrimental for the vaccine efficacy, can potentially be leveraged to identify new protective antigens.^16^ Briefly, allele-specific efficacy, by preferentially preventing infection by pathogen strains with a vaccine-like genotype (which renders those parasites immunologically identical to the vaccine), provides a selective advantage to strains that escape vaccine-induced protection.

Under this scenario, in placebo-controlled vaccine efficacy field trials, breakthrough infections in vaccinees should differ in allele frequency distribution relative to infections in placebo recipients, with the vaccine allele significantly under-represented among vaccinee infections, which can be formally tested with a sieve analysis.^16^ By extension, in the context of a whole organism-based vaccine, a sieve analysis across the whole genome (SA_WG_) performed on parasite isolates collected from participants in randomized, placebo-controlled, field efficacy trials, by revealing protein-coding loci in which the vaccine allele is under-represented in vaccinees relative to controls, may enable the identification of protective targets. These field trials need to meet several conditions; in particular, i) they need to be conducted in a geographic region of high disease transmission, such that genetic diversity in the parasite population is high; ii) the pathogen should have a high rate of effective recombination, such that genetic linkage is limited to only a few hundred base pairs, and consecutive loci are essentially independent; and (iii) vaccine efficacy (VE) is genotype-specific.

Two such clinical trials were completed in regions with intense, seasonal malaria transmission of sub-Saharan Africa. One assessed the efficacy of PfSPZ Vaccine, a radiation-attenuated whole sporozoite vaccine *versus* saline placebo in 80 adults living in Burkina Faso, West Africa, who were cleared of parasites pre-vaccination.^9^ Efficacy against infection was determined by thick blood smear microscopy during illness and every 4 weeks for two malaria seasons (72 weeks total). VE (1 – hazard ratio) was 48% and 46% after 6 and 18 months of follow-up, respectively (*p*-value = 0.061 and 0.018). A second study tested PfSPZ-CVac (CQ),^17^ a chemoattenuated PfSPZ vaccine in which live, infectious PfSPZ were administered to participants receiving chloroquine to eliminate blood stage infection. Participants were not cleared of parasites before vaccination. VE in this study was assessed by thick blood smear during illness and every 4 weeks for a single malaria transmission season (24 weeks). VE compared to saline placebo in 62 participants in this study was 34% (*p*-value = 0.21). Both studies randomized participants 1:1 to receive either the malaria vaccine or placebo. The PfSPZ in both vaccines were from the PfNF54 isolate, the parent stock from which the reference 3D7 was cloned.^18^ In protein-coding regions, PfNF54 and 3D7 differ in fewer than 100 non-synonymous sites, mostly in members of multigene families.^19^

Here, we report the outcome of SA_WG_ performed on Pf isolates collected from vaccinees and control subjects (placebo recipients) from these two vaccine clinical trials when they first became positive, two or more weeks post vaccination. We identified, in each trial, a set of parasite genes in which vaccine genotypes were significantly under-represented in infections from vaccinees relative to controls and prioritize genes identified in both studies. We describe these genes and show enrichment for features expected from protective antigens, consistent with identification of true protective candidates by SA_WG_.

## RESULTS

### Whole genome sequence data and complexity of infection for Pf isolates

Thirty-three participants (13 and 20 in the vaccine and placebo study arms, respectively) in the Burkinabe PfSPZ Vaccine trial were infected at least once with Pf during the follow up period starting two weeks after the last vaccination dose. Total DNA was isolated for each participant’s first infection. An average of 43 million Illumina reads were generated per sample (**table S1**). On average, 18 million reads per sample mapped to the parasite genome (range: 65,600 – 30,890,811). Two samples, one each in the vaccine and placebo arms did not reach the desired coverage threshold and were not used further. Complexity of infection, measured by number of clones per infection or by *F*_WS_, did not differ significantly between study arms (**fig. S1)**. Most samples were polyclonal in both study arms, as expected during the malaria transmission season in a high transmission area; average number of clones per infection was 2.6 and 2.7 in vaccine and placebo recipients, respectively. In the Malian PfSPZ-CVac (CQ) trial, 40 participants (17 vaccinees and 23 who received a placebo) experienced one or more Pf infections during follow up. An average of 60·5 million sequence reads were obtained per sample, and 23 million mapped to the parasite genome (range: 85,469 – 43,912,950) (**table S1**). Four samples from vaccinees and five from placebo recipients did not pass the coverage threshold; 13 and 18 isolates from vaccinees and placebo recipients, respectively, were used in downstream analyses. Once more, complexity of infection did not differ significantly between study arms (**fig. S2)**, and the average number of clones per infection was 3.5 and 3.1 in vaccinees and controls, respectively. A total of 31,122 and 31,659 SNPs in the Pf nuclear genome passed quality filters, respectively, for the Burkinabe and Malian samples (**table S2**) and were used for downstream analyses. As expected for endemic regions of high malaria transmission, the rate of effective recombination in the Burkinabe and Malian Pf populations was high, with linkage disequilibrium between variable sites decaying rapidly to background levels after ∼800 bp in both data sets (**fig. S3**).

### Identification of target sites and target loci with SA_WG_

Since PfSPZ Vaccine efficacy is known to be genotype-specific under certain protocols,^15^ we conducted a whole-genome sieve analysis (SA_WG_) on Pf isolates from each trial, to identify parasite proteins that are putative targets of a protective immune response. A SA_WG_, by revealing genomic sites in which the vaccine allele (here, the allele in PfNF54) is significantly under- represented among vaccinee infections, identifies these “putative protective sites”, or “target sites”.^16,20^ This relies on the rationale that parasites with a vaccine-like genotype at these sites were unable to break through the vaccine-induced protection. Also, we term “target loci” the protein-coding genes that contain non-synonymous target sites, since they encode proteins that are, potentially, the target of a protective response, in a variant-specific manner.

Among the samples from the PfSPZ Vaccine trial in Burkina Faso, 684 significantly differentiated SNPs were identified between vaccinee and placebo recipient Pf isolates, 508 of which were target sites (**table 1**). Of those, 202 were non-synonymous SNPs, which mapped to 165 protein-coding loci or “target loci” (**table 1; table S3**). These loci included several that encode well-established sporozoite antigens, most notably PF3D7_0304600, which encodes CSP, the protein used in the formulation of the RTS,S|AS01 and R21|Matrix-M vaccines, and PF3D7_1335900, which encodes the thrombospondin-related anonymous protein (PfSSP2|TRAP). These are two of the most abundant proteins on the sporozoite surface.^21^ In Mali, 314 target sites were identified. Among those were 110 non-synonymous target sites, mapping to 93 target loci (**table 1; table S4**), including sporozoite antigens PfSSP2|TRAP and E140 (encoded by PF3D7_0104100), among others. The target sites identified in each SA_WG_ were distributed across all 14 chromosomes (**fig. 1**). Some are unique to each study, while other target sites are identified in both studies (**fig. S4**).

**Figure 1.**
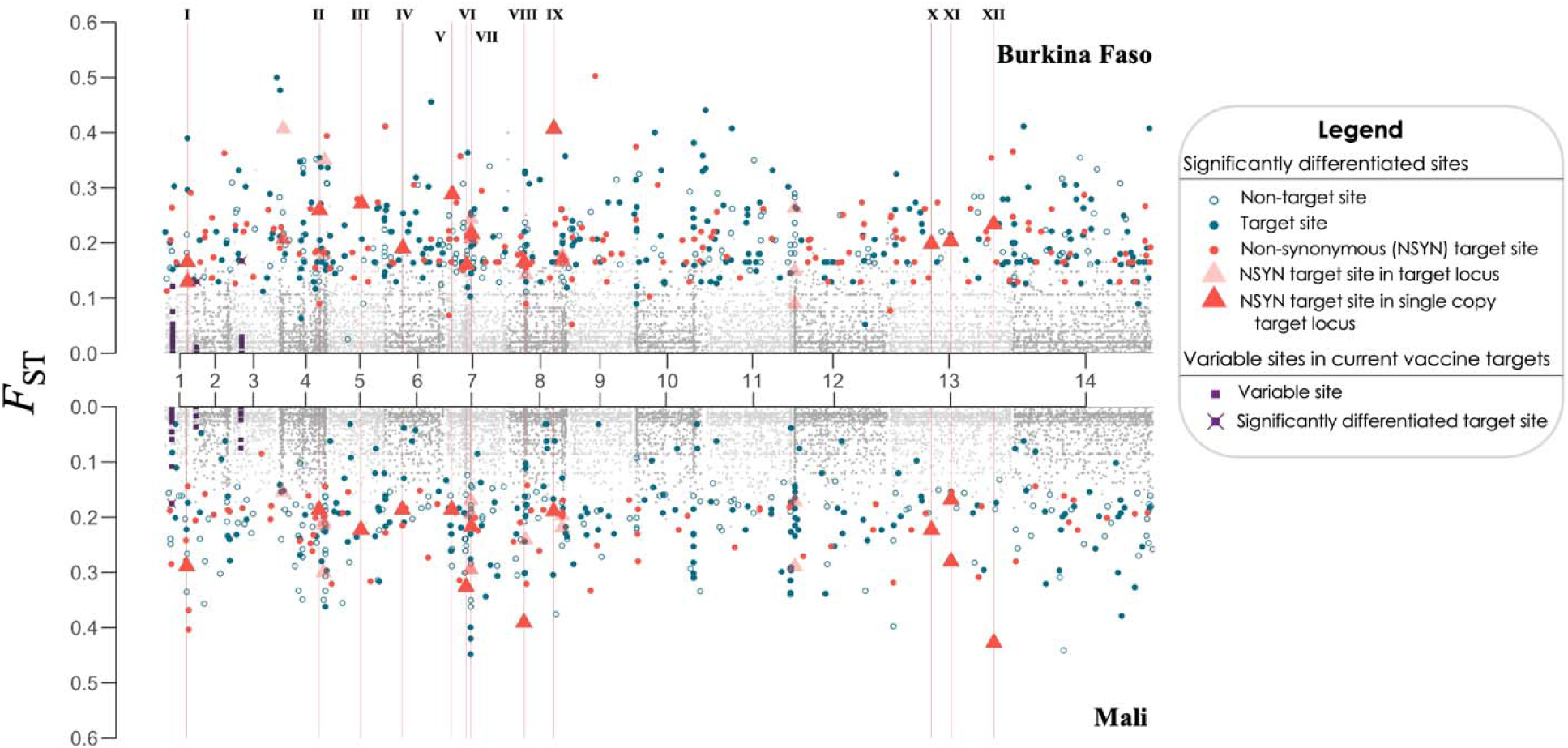
Whole-genome sieve analysis of *P. falciparum* isolates from field trials of two PfSPZ vaccines, in Burkina Faso and in Mali. Site-wise *F*_ST_ values between placebo and vaccinee parasites are shown for Burkina Faso and Mali vaccine trials, separately. Significantly differentiated genomic sites (permutation method; k=5000; *p-*value<0.05) are shown for each trial (non-grey), including sites where the vaccine allele was underrepresented in isolates in controls (open circles) and those where the vaccine allele was underrepresented among vaccinee isolates (target sites; filled circles and triangles). Non-synonymous target sites (red colors) are further highlighted when they were present in the same target locus in both studies (triangles), and the locus is a member of a multigene family (light red triangle); and when they are observed in same single-copy target locus in both studies (dark red triangle). The latter (n=12; shown in roman numerals) are shown across the genome (red, vertical line): PF3D7_0113800 (I), PF3D7_0421700 (II), PF3D7_0518700 (III), PF3D7_0609600 (IV), PF3D7_0703900 (V), PF3D7_0711200 (VI), PF3D7_0713600 (VII), PF3D7_0808100 (VIII), PF3D7_0826000 (IX), PF3D7_1324300 (X), PF3D7_1335900 (XI), PF3D7_1361800 (XII). Variable sites in E140 (in chromosome 1), LSAP2 (in chromosome 2) and CSP (in chromosome 3) are shown (purple). In each of these three loci, one non-synonymous target site is present in one study but not the other.

**Table 1.** Classification of significantly differentiated sites between samples from vaccinees and placebo-recipients. . The country and vaccine used in each case are mentioned in the header, for each study. Significantly differentiated sites in which the vaccine allele was underrepresented among vaccinee samples are termed “target sites” (dark blue columns); the loci to which non-synonymous (NSYN) target sites (tan) map are termed “target loci” (brown).

|  | Burkina Faso, PfSPZ Vaccine trial |  | Mali, PfSPZ CVac trial |  |
| --- | --- | --- | --- | --- |
|  | Vaccine allele depleted in vaccinee samples (target sites and target loci) | PfNF54 allele depleted in placebo recipient samples | Vaccine allele depleted in vaccinee samples (target sites and target loci) | PfNF54 allele depleted in placebo recipient samples |
| **Significantly differentiated sites** | 508 | 176 | 314 | 196 |
| **Synonymous (SYN)** | 94 | 34 | 47 | 47 |
| **Non-Synonymous (NSYN)** | 202 | 60 | 110 | 69 |
| **Other (intergenic, etc.)** | 212 | 82 | 157 | 80 |
| **Protein-coding loci with differentiated NSYN sites** | 165 | 48 | 93 | 58 |

Several factors can lead to potential false positive signals in SA_WG_ (i.e., polymorphic sites identified as “target sites” but that are not the target of a protective response), including differential genotype assortment into study arms by chance (a common challenge for studies with relatively small sample size, such as the two studies here) and genomic sites in linkage disequilibrium with protective sites.^16,20^ These factors can also explain the existence of significantly underrepresented vaccine alleles in placebo recipients (**table 1**, blue columns; **fig. 1**, open blue dots).

To determine if these SA_WG_ were likely to have revealed functionally protective proteins, we sought to determine whether the target sites and associated target loci displayed patterns expected from antigens. Several lines of evidence suggest this to be the case, as follows.

Analyses of the 262 non-synonymous sites significantly differentiated between vaccinee and control infections in the SA_WG_ of Burkinabe samples revealed that the vaccine allele was significantly depleted primarily among samples from vaccine recipients (n = 202; 77.1%) *versus* placebo recipients (n = 60; 22.9%) (**table 1**; **table S5**; *p*-value < 0.001), as expected from the genotype-specific sieve effect of a vaccine-primed immune system. In the Malian study, the vaccine allele was again significantly depleted more frequently among samples from vaccine (61.5%) *versus* placebo recipients (38.5%) (**table 1**; **table S5**; *p*-value < 0.001). Additionally, a GO term enrichment analysis with the 165 target loci identified with the SA_WG_ from the Burkinabe clinical trial samples revealed a significant enrichment of membrane-associated proteins and proteins involved in host-parasite interactions, traits common among antigens (**fig. S5**). A similar analysis of the 93 target loci identified in the analysis of the Malian samples (a study in which VE was not significant) did not reveal GO term enrichment. Enrichment analyses on the loci with significantly differentiated non-synonymous SNPs in which the vaccine allele was depleted in controls did not show significant enrichment of GO categories in either study (**fig. S5**), consistent with the presence of many false positives among these gene sets.

Possibly the most compelling lines of evidence that SA_WG_ identifies *bona fide* protective targets rest with the number and nature of the target loci that were identified in both studies, and in the fact that target sites are immunologically relevant. These aspects are detailed in the next two sections.

### Twelve single-copy target loci were common to both studies

Among all target loci identified in each study, 18 were common to both studies (**table 2**), a set larger than expected by chance (*E* = 5.0; *p-*value ∼3.7x10^-7^), given the number of loci with non-synonymous variable SNPs across studies (n = 3326; **table S2)**. In fact, the observed value of 18 shared target loci was found outside of the 99% confidence interval of the expectation (99%CI = [0, 10]). Among these 18 target loci, twelve are single copy genes and the remaining six are members of multigene families. Since SA_WG_ false positives could arise from read mapping artifacts among members of multigene families, we focused further on single copy genes. Given 138 and 80 single copy target loci in the Burkina Faso and Mali studies, respectively, and a total of 2848 single copy genes with non-synonymous variable sites, the probability of finding twelve single copy target loci in common between studies is also significantly higher than expected (*E* = 3.9, 99%CI = [0, 9]; *p-*value ∼3.6x10^-4^). Finally, as expected for protective targets of pre-erythrocytic vaccines, transcripts for all twelve single-copy target loci are present in sporozoites, and again during the liver stage, although one is only detected by day 6 (**table 2**).^22,23^ Encouragingly, one of the targets common to both studies is PfSSP2|TRAP, a well-established pre-erythrocytic antigen, as well as several conserved, membrane-associated Pf genes encoding proteins of unknown function, possibly novel candidate vaccine targets.

**Table 2.**
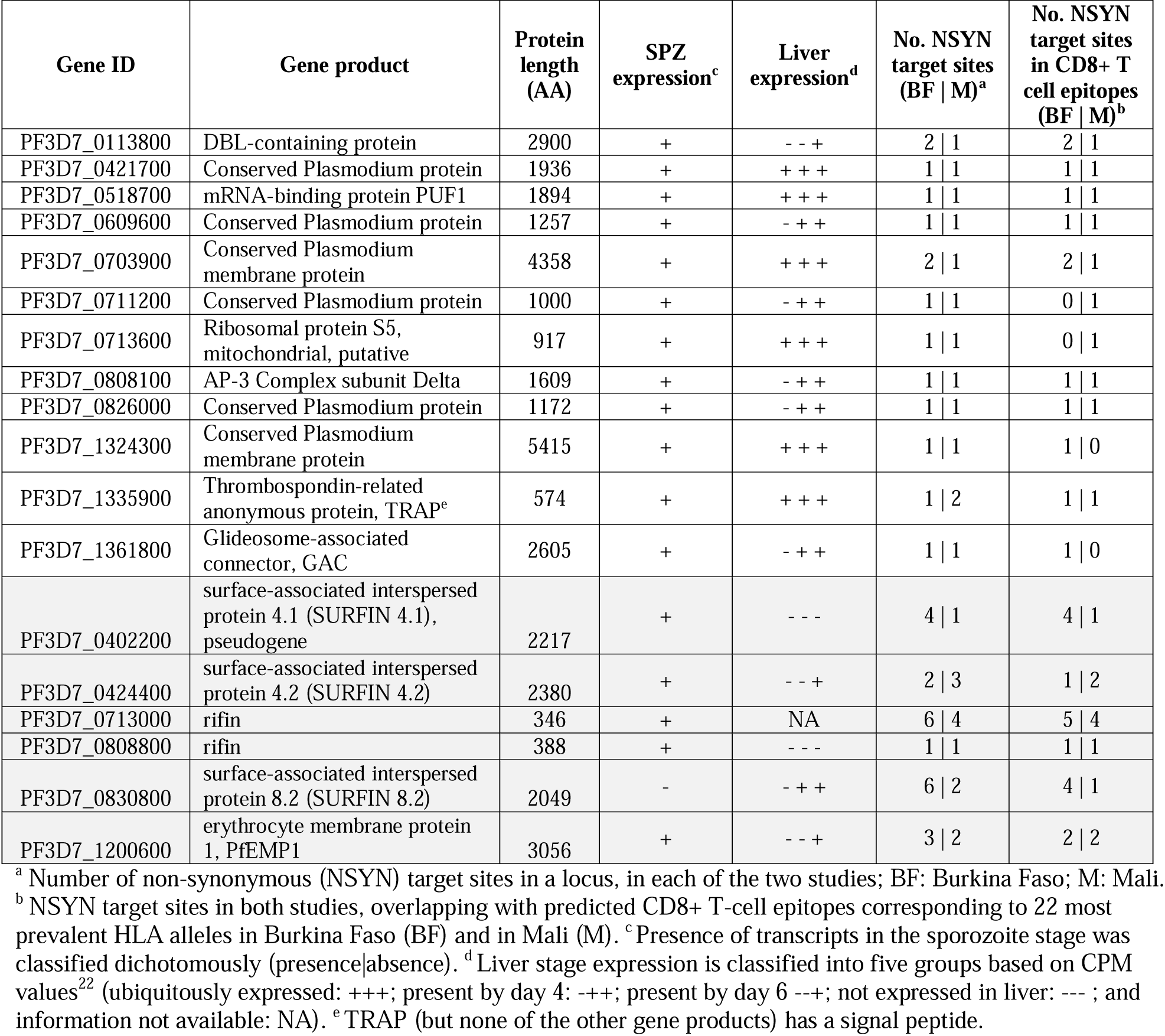
Target loci identified in both studies. The 18 target loci are shown, each containing at least one non-synonymous (NSYN) target site (i.e., significantly differentiated NSYN SNPs between samples from vaccinees and controls, with the vaccine allele depleted in the former) in both the studies and associated gene expression. Single-copy genes (not shaded) and members of multigene families (shaded rows) are shown.

### Target sites are in immunologically relevant regions of target loci

CD8^+^ T cells play a critical role in protection following immunization with Pf sporozoites (SPZ).^24–26^ Therefore, evasion of PfSPZ vaccine-induced protection could be associated with polymorphisms in CD8^+^ T cell-restricted epitopes present in Pf SPZ or liver stage antigens that prevent parasite detection by T cells primed with the Pf NF54 (vaccine) variant, in protective antigen-encoding loci. Accordingly, to determine if the target sites identified with SA_WG_ were immunologically relevant, we predicted the CD8^+^ T cell immunopeptidome of Pf NF54. Since the HLA genotype of individual participants is not available, we defined the CD8 T cell immunopeptidome of Pf NF54 as the union of all non-synonymous sites in NF54 genomic sequences that encode predicted CD8^+^ T cell strong-binding epitopes restricted to the 22 most common HLA alleles in Mali and Burkina Faso. We observed that, in fact, most target sites fall within the CD8^+^ T immunopeptidome (**table 2**), even though enrichment is not significant (**table S6**). The need to use a large HLA allele set to define the immunopeptidome resulted in inclusion of roughly 3/4 of all amino acid residues in the immunopeptidome, possibly reducing statistical power (**table S6**).

We wanted to compare the distribution of target sites in current malaria vaccine pre-erythrocytic targets relative to the immunopeptidome. The vaccine development pipeline for malaria includes twelve pre-erythrocytic antigens, including CSP (PF3D7_0304600) and PfSSP2|TRAP (PF3D7_1335900).^27^ We found 138 and 135 variable sites among these twelve antigens in Burkinabe and Malian sample sets, respectively, but only a small subset of those are target sites (**tables S3** and **S4**). Among samples from Burkina Faso, three of the 202 non-synonymous target sites mapped to three of those twelve antigens, one each in CSP, PfSSP2|TRAP, and LSAP2 (PF3D7_0202100) (**fig. 1; table S3**). Importantly in the context of the current WHO-endorsed malaria vaccines, we note that the target site in CSP (polymorphism in Pf3D7 CSP nucleotide 970 C>A, leading to a Gln324Lys change) falls in the Th2R region, which contains several MHC Class I-restricted epitopes.^28^ Among Malian samples, two target sites were identified among these twelve vaccine antigens, both of which map to PfSSP2|TRAP (**table S4**).

PfSSP2|TRAP presents as a particularly relevant case study, given its status as a pre-erythrocytic vaccine candidate, and its identification as a target locus in both SA_WG_ studies, since the vaccine allelic state in three non-synonymous, polymorphic site were significantly under- represented in vaccinees relative to controls (**fig. S6**). Analysis of the distribution of target sites relative to predicted immunopeptidome reveals that the target site identified in the Burkinabe study (829 T>A, leading to Leu277Ile) falls within the predicted and/or validated CD8+ T cell immunopeptidome, as does one of the target sites identified in the Malian study (542 G>T, leading to Arg181Leu) (**table 2**, **fig. 2**). To investigate the possible contribution of CD8+ T cells to the reduction in frequency of parasites carrying the vaccine allele in vaccinee infections, we considered all peptide variants identified in each study (residue containing the target site plus the eleven residues up- and down-stream from it) and the distribution within them of predicted CD8 T cell epitopes. In the case of target sites in residues 66 and 181, the vaccine peptide variant was the most common variant across all infections, while the target site in residue 277 was the third most common in the Burkinabe study and was rare in Mali (**fig. S7**). In each case, the vaccine variant was more common in infections from controls than from vaccinees, but lack of power from small sample size (and the low frequency of the vaccine variant) precludes significance testing (**fig. S7**). Interestingly, in the Burkinabe study, where the vaccine variant in residue 277 is common to two of the seven variants detected in that study and only that variant is recognized by a common HLA allele, the vaccine variant is under-represented in vaccinee infections relative to those in controls (**fig. 2**), suggesting that recognition, by CD8^+^ T cells, of cells infected with Pf this genotype could in fact contribute to their elimination from the vaccine arm.

**Figure 2.**
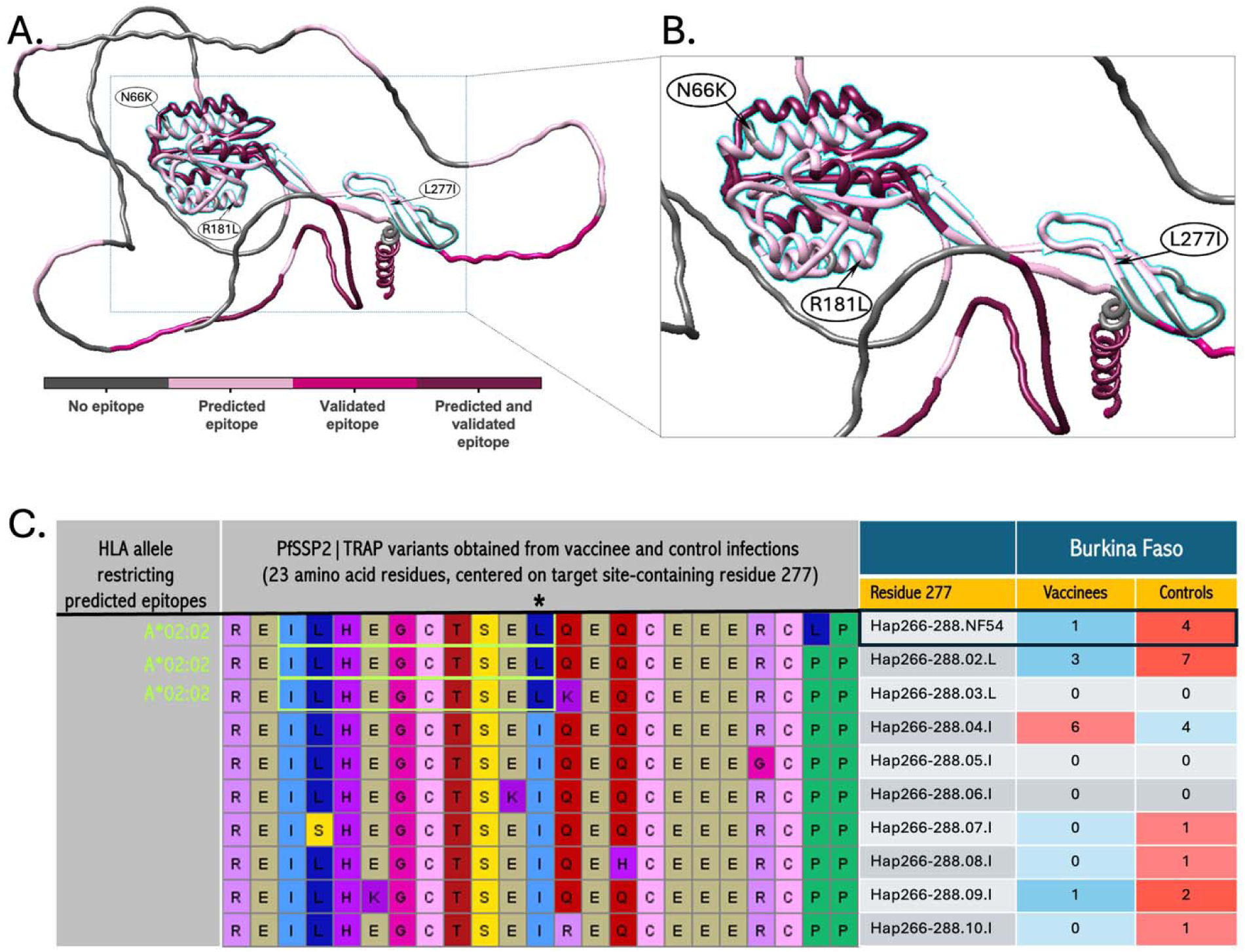
Genetic variation and distribution of predicted CD8^+^ T-cell epitopes at PF3D7_1335900, the locus encoding PfSSP2|TRAP. **A.** 3D structure of TRAP, obtained with AlphaFold.^31^ Amino acid residues containing target sites are noted, with residue coordinate (L66I; R181L; L277I). Magenta color spectrum shows protein regions with predicted strong binding CD8^+^ T-cell epitopes, experimentally validated epitopes or both. Display generated with UCSF Chimera v1.15.^32^ **B.** Magnification of protein regions containing target sites. Shading (in light blue) encapsulates structurally important motifs required for adhesion and invasion of host cells, including a von Willebrand factor A (vWA)-like fold (top left)^29^ and a thrombospondin type 1 repeat (TSR) domain (center right).^30^ **C.** Alignment of TRAP peptide sequences at and surrounding residue 277 (central panel; amino acid residues 266-288), with location of epitope restricted to HLA A*02:02 marked (green box). Frequency of each variant identified in Pf infections from vaccinees or controls is shown (right panel). Vaccine peptide in black frame. Color-coding: same color in both arms when a variant was found in equal number in vaccinees and controls; strong red/blue when frequence was double of more (red) in one arm than the other (blue); light blue/red otherwise.

All three target sites found in PfSSP2|TRAP, including the site in the codon encoding the variant Asn66Lys (target site 198 C>G), fall within the two adhesive domains required for host cell adhesion (**fig. 2**).^29,30^

## DISCUSSION

In 2001, Gilbert and colleagues^33^ proposed an approach they termed “sieve analysis” to determine if VE is allele(genotype)-specific, and to quantify the relative risk of infection, post-vaccination, associated with strains encoding different variants of a vaccine immunogen. Our group conducted the first comprehensive sieve analysis in malaria to identify residues and epitopes associated with protection post-vaccination with the apical membrane antigen 1 (AMA1) variant encoded by 3D7, and in repeat infections after natural exposure to Pf malaria, and identified several key residues important for protection.^34–36^ More recently, we conducted a genome-wide comparative study of genetic variation between groups of individuals with differing levels of acquired immunity, to identify potential Pf protein targets of acquired immunity to clinical malaria.^37^ Merging the concepts of sieve analysis and genome-wide differentiation studies between Pf genomes from phenotypically different groups of individuals, we recently proposed SA_WG_, an extension of the sieve analysis concept to the whole genome of a genetically diverse, recombining parasite species, such as Pf, using samples from a placebo-controlled vaccine trial; by detecting genomic sites in which the vaccine allele is significantly under-represented in vaccinee Pf infections relative to those from placebo recipients, a SA_WG_ should thereby identify targets of vaccine-induced protection.^16^ Here, we conducted the first SA_WG_ on Pf isolates collected from VE trials of PfSPZ vaccines conducted in malaria-endemic areas, with the explicit purpose of identifying potential targets of PfSPZ vaccine-induced protection. The SA_WG_ conducted on the Burkinabe study isolates revealed 165 target loci (138 of which are single-copy genes) while the Malian SA_WG_ revealed 93 target loci (80 single-copy genes). A total of 18 target loci were identified in both analyses, including twelve single-copy genes, numbers significantly higher than expected by chance.

Other lines of evidence are consistent with the possibility that several of these twelve target loci may be targets of PfSPZ vaccine-induced protection. Most notably, on of twelve single copy targets, PF3D7_1335900, encodes one of the most well-established pre-erythrocytic sporozoite stage antigens, PfSSP2|TRAP.^38^ Also, PF3D7_0113800 encodes a DBL domain known to activate Rho GTPases involved in hepatocyte invasion during Pf infection,^39,40^ and PF3D7_1361800 encodes the glideosome-associated connector (GAC), a crucial component of the gliding machinery that enables host tissue transversal, cell invasion and egress.^41^ GAC connects actin fibers to surface adhesins (including TRAP), playing a pivotal role in parasite forward movement. Interestingly, GAC becomes exposed and is shed as parasites migrate through host tissue,^41^ providing an opportunity for immune system priming and subsequent parasite detection. PF3D7_0703900 and PF3D7_1324300 encode conserved, membrane-associated proteins of unknown function, but may be accessible to either cell-mediated or humoral immunity if present on the sporozoite surface. PF3D7_0518700 encodes the mRNA-binding protein PUF1, which may regulate stability and translation efficiency of specific mRNAs important for gametocyte development.^42^ PF3D7_0808100 encodes AP-3 complex subunit delta, with a role in endosomal protein trafficking and vesicle formation within the parasite.^43,44^ PF3D7_0713600 encodes a putative mitochondrial ribosomal protein S5. Even though the relationship between some genes identified by SA_WG_ and vaccine-induced protection is not immediately clear from the localization of the encoded proteins, surface exposure of targeted proteins is not a requirement for CD8^+^ T cell-effected protection. The SA_WG_ revealed four additional parasite genes encoding conserved proteins of uncharacterized function (**Table 2**).

One of these, PF3D7_0421700 has been identified as a likely target of naturally acquired immunity to clinical malaria,^37^ suggesting an overlap between protective targets of sporozoite-based vaccination and natural infection. Finally, within these 12 target loci, most non-synonymous target sites are located within predicted CD8^+^ T cell epitopes, consistent with cell-mediated immunity (**Table 2; table S4**).

The two SA_WG_ yielded different numbers of target loci, which partially overlapped between studies. Differences can result from several factors, including Pf treatment (or lack thereof) before vaccination, the vaccine tested, and VE outcome. The Burkinabe clinical trial^9^ tested three doses of PfSPZ Vaccine (2.7x10^6^ PfSPZ|dose), which consists of radiation attenuated, non-replicating PfSPZ that arrest early in the liver stage. The Malian trial^17^ assessed three doses of PfSPZ-CVac (CQ) (2x10^5^ PfSPZ|dose), which consists of non-attenuated, fully replicating PfSPZ that are eliminated by chloroquine after liver schizonts rupture and parasites reach the blood.^5^ Therefore, different antigen sets are presented by the two vaccines, and for different periods of time, likely inducing responses to different, and only partly overlapping, sets of targets. Patients were treated to clear blood stage parasitemia pre-vaccination in the Burkinabe study, but not in Mali. It is unclear if an ongoing infection with a genotype different from PfNF54 could have impacted protection outcome in a genotype-specific manner, hence contributing to an increase in noise to signal ratio, with a larger proportion of the differentially distributed genotypes being due to chance in the Mali study. In addition, the two studies differed in protection outcome, with the trial in Burkina Faso (but not the Malian trial) resulting in significant VE. Lack of significant VE likely also increases noise to signal ratio. This is consistent with the lower signal observed in site and gene enrichment analyses in Malian relative to Burkinabe samples, including a smaller proportion of differentially distributed non-synonymous sites in which the vaccine allele was underrepresented among vaccinee samples relative to controls, and a lack of GO term enrichment among target loci. Therefore, the studies almost certainly differ in power to detect an association between Pf genetic variants and protection. Differences between host populations in the two studies, including in HLA allele frequencies, previous malaria exposure, and previous exposure to different Pf genotypes, could also have led to differences in the target sites detected in these two small trials.

This study has two key limitations: one was the small sample size of each SA_WG_, which likely contributed to increase the number of false positives;^16^ the second was the unavailability of HLA genotype for each participant. The lack of this information resulted in reduced power to detect an associated between target sites and individual immunopeptidomes, and precluded paired host HLA-Pf genotype analysis to determine the extent to which HLA genotype contributed to Pf genotype differentiation between vaccinees and controls. Finally, a limitation inherent to any SA_WG_ is the inability to identify sites that encode residues targeted by, and which contribute partially to, the protective immune response but that were invariable within the pathogen data set collected. Without genetic variation, it is impossible to detect pathogen genotype differentiation between study arms, the metric upon which SA_WG_ relies.

Overall, the encouraging results obtained support the use of SA_WG_ as an agnostic approach to identify novel targets of vaccine-induced protection. Analyses of additional PfSPZ-based placebo-controlled VE trials in malaria-endemic areas can be used to further test this approach, potentially strengthening identification and prioritization of target loci. Importantly, it is essential to validate these results for any targets intended for vaccine formulation, by demonstrating that they are both immunogenic and protective. Finally, the evidence presented is consistent with a scenario where multiple parasite proteins contribute to the protective immune response induced by PfSPZ vaccines and in which these protective immune responses are often allele-specific, as is the case, by definition, of the target loci identified here. Therefore, to achieve strain-transcendent, broad protection, PfSPZ vaccines will need to induce optimal protective immune responses to variable epitopes from multiple proteins, including subdominant epitopes, and could potentially be improved by including mixtures of immunologically complementary PfSPZ strains, genetic crosses of parasites from different geographic regions, and|or co-expression, in an individual transgenic Pf vaccine strain, of multiple, immunologically complementary variants of target antigens.

## RESOURCE AVAILABILITY

Whole genome shotgun sequencing data are available under BioProject PRJNA1134691 (BioSamples SAMN43081578-SAMN43081613) for Burkinabe samples and BioProject PRJNA1134692 (BioSamples SAMN43082681 - SAMN43082748) for Malian samples. In-house scripts can be found at https:||github.com|igs-jcsilva-lab|.

## Data Availability

Whole genome shotgun sequencing data are available under BioProject PRJNA1134691 (BioSamples SAMN43081578- SAMN43081613) for Plasmodium falciparum samples from Burkina Faso, and BioProject PRJNA1134692 (BioSamples SAMN43082681 - SAMN43082748) for Plasmodium falciparum Malian samples. In-house scripts can be found at https:||github.com|igs-jcsilva-lab|.

## ACKNOWLEDGEMENTS

The authors thank Maryland Genomics for the preparation of all genomic libraries and generation of whole genome sequencing data. We thank Drs. Gloria Meng-Hsuan Lin and Dominique Soldati-Favre for discussions on the GAC protein presentation. We further thank the volunteers and their families in Mali and Burkina Faso for their participation in clinical trials of PfSPZ Vaccine and PfSPZ-CVac.

## AUTHOR CONTRIBUTIONS

Conceptualization: JCS, AD, STH, SLH

Sample acquisition: BS, SJ, AO, ABT, DC, MAT, SBS, MBL

Methodology: JCS, AD, DGH, RJS

Data Analysis: AD, RJS, DGH, TCS, OOI

Data Management: CG, JBM

Visualization: AD, RJS

Funding acquisition: JCS, STH, MBL, CVP

Project administration: JCS

Supervision: JCS

Writing – original draft: JCS, MBL, AD, RJS

Writing – review & editing: All co-authors

## MATERIALS AND METHODS

### Study design

This study aimed to identify parasite proteins putatively targeted be the immune system in response to vaccination with whole Pf sporozoites (PfSPZ) using whole genome sieve analysis (SA_WG_). To this end, for two placebo-controlled malaria vaccine efficacy field trials of PfSPZ-based vaccines, we compared allele frequency distribution, at each genomic site, between Pf infections collected from the unprotected vaccinees and from placebo recipients. In each study, we identified genomic sites in which the vaccine allele (allelic state in PfNF54) was significantly under-represented among vaccinee infections, relative to infections from control individuals.

### Samples

Samples originated from two clinical trials assessing the efficacy of malaria vaccines against natural infection in West Africa, one in Burkina Faso (NCT02663700) and one in Mali (NCT02996695). Samples analyzed correspond to the timing of the first thick blood smear-positive malaria infection occurring at least two weeks after the last vaccination dose, corresponding to 33 and 40 participants, respectively, in the Burkinabe and Malian trials.

Samples consisted of a 2 mL venous blood draw that was leukocyte-depleted, pelleted and stored at -20°C or -80°C. DNA was extracted from each sample, using the Qiagen Blood DNA Midi Kit (Valencia, CA, USA). Total DNA extracted varied between 8.0-1031.2 ηg (median: 75.9 ηg) for Burkinabe samples and between 0.0-78043.3 ηg (median: 10.3 ηg) for Mali samples. Nine out of 40 samples from Mali had unusually high DNA amounts, likely due to contamination. All samples with <100 ηg of total DNA or those with likely contamination underwent selective whole genome amplification (sWGA) with Pf-specific primers to increase the relative amount of parasite DNA. Amplification was conducted in a 0.2 mL 96-well PCR plate. The reaction mixture comprised 1X bovine serum albumin (BSA), 1 mM deoxynucleotide triphosphates (dNTPs), 2.5 µM of each amplification primer, 1X Phi29 reaction buffer, and 30 units of Phi29 DNA polymerase. A total of 17 µL of template DNA was added to the reaction mixture, bringing the final reaction volume to 50 µL. The amplification was carried out in a thermocycler using a step-down protocol: 35 °C for 5 min, 34 °C for 10 min, 33 °C for 15 min, 32 °C for 20 min, 31 °C for 30 min, and 30 °C for 16 h. Enzyme inactivation was achieved by heating at 65 °C, followed by cooling at 4 °C.^45^ Three out of 40 Malian samples had insufficient DNA for library construction (**table S1**).

### DNA sequencing

Genomic DNA, extracted from leukocyte-depleted samples or generated through sWGA, was used to construct DNA libraries using the KAPA Library Preparation Kit (Kapa Biosystems, Woburn, MA). DNA was fragmented with the Covaris E220 to ∼200-300 bp. Libraries were prepared using a modified version of manufacturer’s protocol. The DNA was purified between enzymatic reactions and library insert size selection was performed with AMPure XT beads (Beckman Coulter Genomics, Danvers, MA). When necessary, a PCR amplification step was performed with primers containing an index sequence of six nucleotides in length. Libraries were assessed for concentration and fragment size using the DNA High Sensitivity Assay on the LabChip GX (Perkin Elmer, Waltham, MA). Library concentrations were also assessed by qPCR using the KAPA Library Quantification Kit (Complete, Universal) (Kapa Biosystems, Woburn, MA). The libraries were multiplex on a 150 bp paired-end Illumina HiSeq 4000 run (Illumina, San Diego, CA). For each sample, sequencing data was generated to a minimum of 75% breadth of coverage of the Pf3D7 genome with at least 5X depth of coverage. Samples for which that level of coverage was unfeasible, due to a high proportion of human DNA, were not used for downstream whole-genome sieve analysis.

### Sequencing variant identification and classification

Reads were aligned to the 3D7 reference genome obtained from PlasmoDBv24, in EuPathDB,^46^ using Bowtie2 v2.3.4.3.^47^ All WGS data files with data from the same original specimen were combined, resulting in a merged BAM file that contained all reads regardless of library type (**table S1**). The resulting BAM files were processed according to GATK’s Best Practices documentation.^48^ Joint SNP calling was done using Haplotype Caller v1.7.0. In addition to 31 samples from Burkina Faso PfSPZ and 31 samples from Mali PfSPZ-CVac trial, short read (150 bp) whole genome shotgun sequencing (WGS) data for 1,289 *P. falciparum* isolates collected from 20 malaria-endemic countries in Africa, Southeast Asia, Oceania and S. America were downloaded from NCBI’s Short Read Archive and were used in joint SNP calling (**table S7**).

Diploid calls were allowed to account for heterozygous positions in polyclonal samples. Additional hard filtering was done to remove potential false positives using filter DP < 5 || QUAL < 50 || FS > 14·5 || MQ < 20. In downstream analysis, only single nucleotide polymorphisms (SNPs) were used, and additional filtering was done on the SNP data. Sites with genotype information missing in >30% of samples were removed, as were sites with variants present in fewer than three samples, to avoid possible artifacts. Genomic sites were classified as intergenic, intronic, synonymous and non-synonymous using snpEFF v4.3t.^49^ For genotyping of loci of interest with potential sequence length variation, an in-house pipeline for *in silico* capture and assembly (ISCA v1.2.3; <u>https:||github.com|igs-jcsilva-lab|ISCA</u>) of short sequence reads that map to each of those loci was used.

### Multiplicity of infection

Multiplicity of infection was assessed with the statistic *F*_WS_, estimated with the R package moimix.^50^ Difference in *F*_WS_ difference between study arms was tested with a Wilcoxon Rank Sum test. The number of clones per infection was estimated with dEploid-IBD v0.5,^51^ with default parameters, and significance tested with a chi-square test.

### Whole genome sieve analysis (SA_WG_)

A SA_WG_ was conducted separately using WGS data generated from isolates collected in each clinical trial, and in each case was based on all polymorphic sites that passed filters, as described above. A SA_WG_ consists of two steps: *i*) identification of parasite genomic sites significantly differentiated between study arms (vaccinees and controls), and (*ii*) identification of sites from (*i*) where the vaccine allele is depleted among vaccinee infections. For each genomic site, difference in allele frequency distribution between study arms (step *i*) was quantified using Wright’s Fixation Index, *F*_ST_ (Weir and Cockerham’s implementation in vcftools v0.1015).^52^ For each analysis, *F*_ST_ value significance was determined by randomization of samples across study arms, with 5000 replicates. Sites found to be significantly differentiated between vaccinee and control infections, and in which the vaccine (PfNF54) allele was underrepresented in vaccinee infections, were termed “*target sites*”. Deviation of expected frequency of target sites per study arm was tested with a chi-square test.

Loci were termed “*target loci*” if they contained non-synonymous target sites. The 99% confidence interval and p-value for the number of target loci found, by chance, in both SA_WG_ (given *m* and *n* targets genes, one set in each study), given the total number of genes *g* with variable non-synonymous sites across studies, were both determined by Fisher’s exact conditional test for 2x2 independence (fixed row and column sums, *m* and *n*), which is a conservative version of a multinomial (unconditional) test. The multinomial test is more appropriate (fixed *g* but variable number of target loci, *m* and *n*) but it is computationally expensive. The expected number of target loci that overlap between studies, *E*, is given by *E* = *m*n*|*g*.

### Feature Identification and enrichment analyses

CD8+ T cell epitopes were predicted across target loci using NetMHCpan v4.1,^53^ using default parameters. Only strong binding epitopes were considered in downstream analysis. The HLA alleles used for epitope prediction were the 22 most common among those identified in Mali and Burkina Faso.^54^ Enrichment analyses of gene ontology (GO) terms among different gene sets were done using ShinyGO v0.741.^55^ In target loci, enrichment of target sites in strong binding epitope-encoding regions was tested using a chi-square test.

