## SupplementalMaterials for "Protective targets of PfSPZ vaccines identified from whole-genome sieve analysis of isolates from malaria vaccine efficacy trials in West Africa"

**Supplemental Figures and Tables**

**Supplemental Figures**

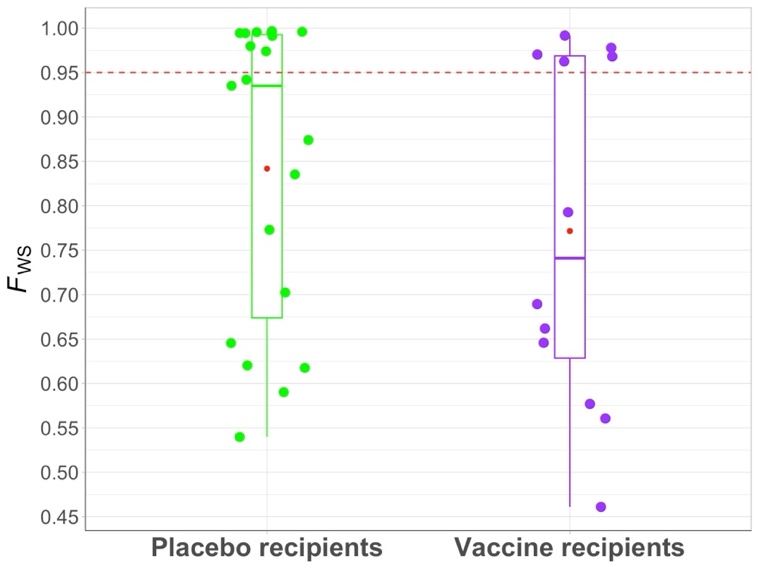

**Figure S1. Multiplicity of infection in samples from the Burkina Faso PfSPZ Vaccine study, as estimated by *F*_WS_.** Boxplot of *F*_WS_ values, with upper and lower quantiles (box edges), median (horizontal line within box) and mean (red dot) shown. *F*_WS_ is a measure of within-host parasite genetic diversity relative to the diversity in the population and was estimated based on bi-allelic sites in the core nuclear genomes of 31 *P. falciparum* samples that passed breadth of coverage cutoff, using R package moimix. The *F*_WS_ values for 19 Placebo and 12 Vaccine recipients are represented by green and purple dots, respectively. The *F*_WS_ = 0.95 threshold (red dotted line) is used to characterize samples as monoclonal (*F*_WS_ ≥ 0.95) and polyclonal (*F*_WS_ < 0.95). The result was corroborated by the estimated number of clones per infection which varied between 2 and 4 in each study arm, with mean of 2.6 among vaccinees (with an average dominant strain proportion of 64%) and 2.7 among placebo recipients (average dominant strain proportion of 61%).

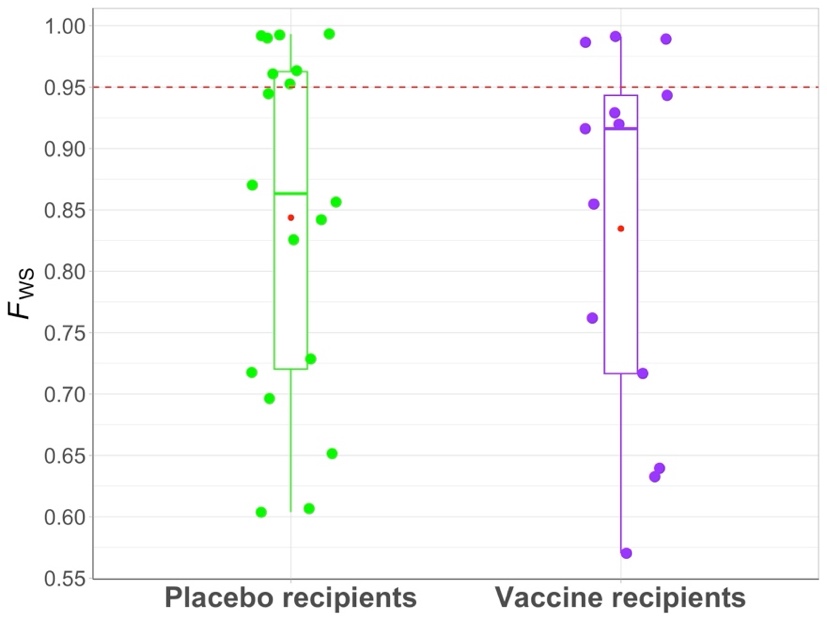

**Figure S2. Multiplicity of infection in samples from the Mali PfSPZ CVac study, as estimated by *F*_WS_.** The *F*_WS_ values for 18 Placebo and 13 Vaccine recipients are represented by green and purple dots, respectively. Details as in Figure S1. There were, on average, 3⋅5 and 3⋅1 clones per infection (average dominant strain proportion of 58% and 59%) among vaccinees and placebo recipients, respectively. In both study arms, the number of clones per infection varied between 2 and 4.

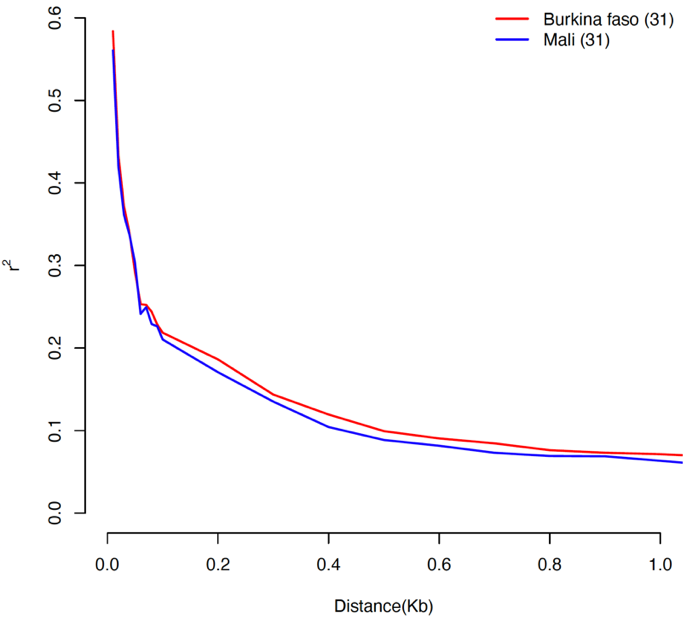

**Figure S3. Distribution of linkage disequilibrium with distance between markers.** Linkage disequilibrium, as measure by *r*^2^, decays rapidly to genomic background levels within 800 bp.

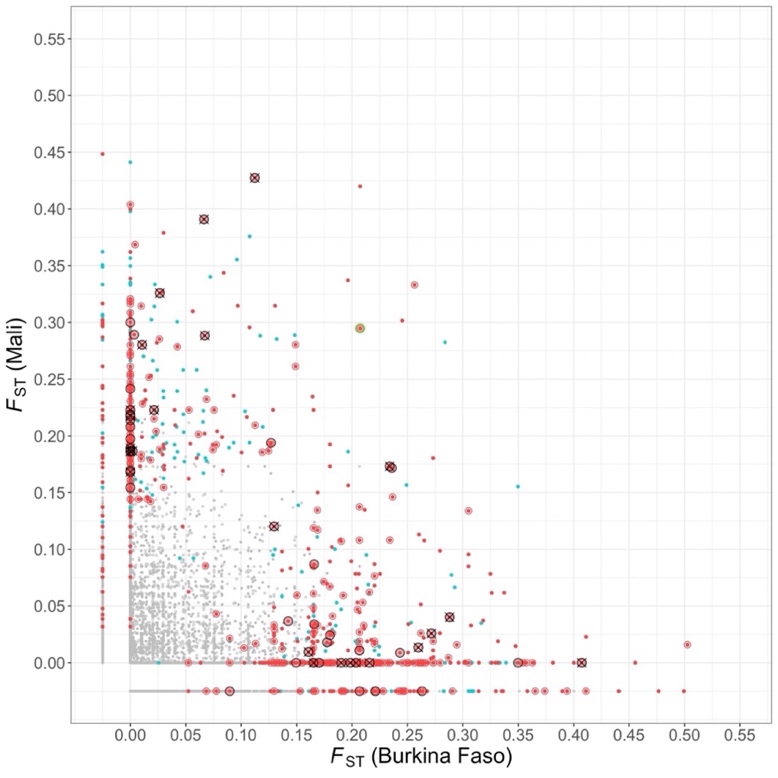

**Figure S4. Target sites identified in the two SA_WG_ from Mali and Burkina Faso vaccine trial samples.** This figure shows the scatterplot of site-wise *F*_ST_ values between placebo and vaccinees groups for both, Mali and Burkina Faso vaccine trials. Significantly (permutation method; k=5000; *p-value*<0.05) differentiated genomic sites are shown for each trial, including sites where the vaccine allele is depleted among vaccinee isolates (red) and those where the vaccine allele is depleted in controls (blue). Among the sites where the vaccine allele is depleted in vaccinee isolates, we further highlighted: non-synonymous sites (concentric red), non-synonymous sites observed in same gene (concentric red circles with a black border) and non-synonymous sites observed in same single-copy gene (concentric red circles with black circle cross) in both the studies are shown. The two same significantly differentiated non-synonymous target sites identified in PF3D7_0713000 (rifin) in both the studies (concentric red circles with a green border) are shown.

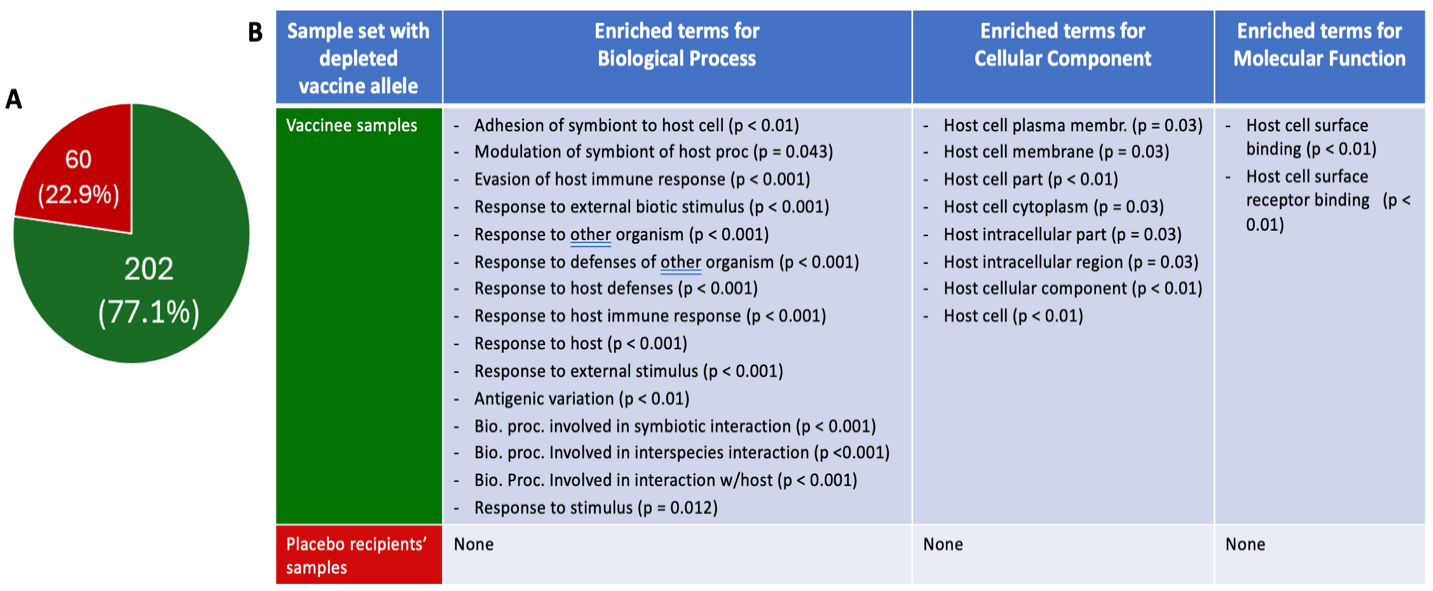

**Figure S5. Distribution of significantly differentiated non-synonymous sites by study arm, and enrichment analyses of target loci.** A) For the Burkina Faso study, significantly differentiated non-synonymous (NSYN) sites were partitioned into two groups: 1) NSYN sites in which the vaccine allelic state was depleted in vaccinee samples, i.e. NSYN target sites (green) and (2) those in which the vaccine allele was depleted in controls (red). B) All unique loci containing these SNPs were used as inputs for two GO term enrichment analyses, using ShinyGO, performed separately for the loci with NSYN sites in (1) and in (2) above. Significance cutoff of false discovery rate, FDR, <0.05 was used. Similar analyses were performed with the Malian data but neither study arm had significant GO term enrichment.

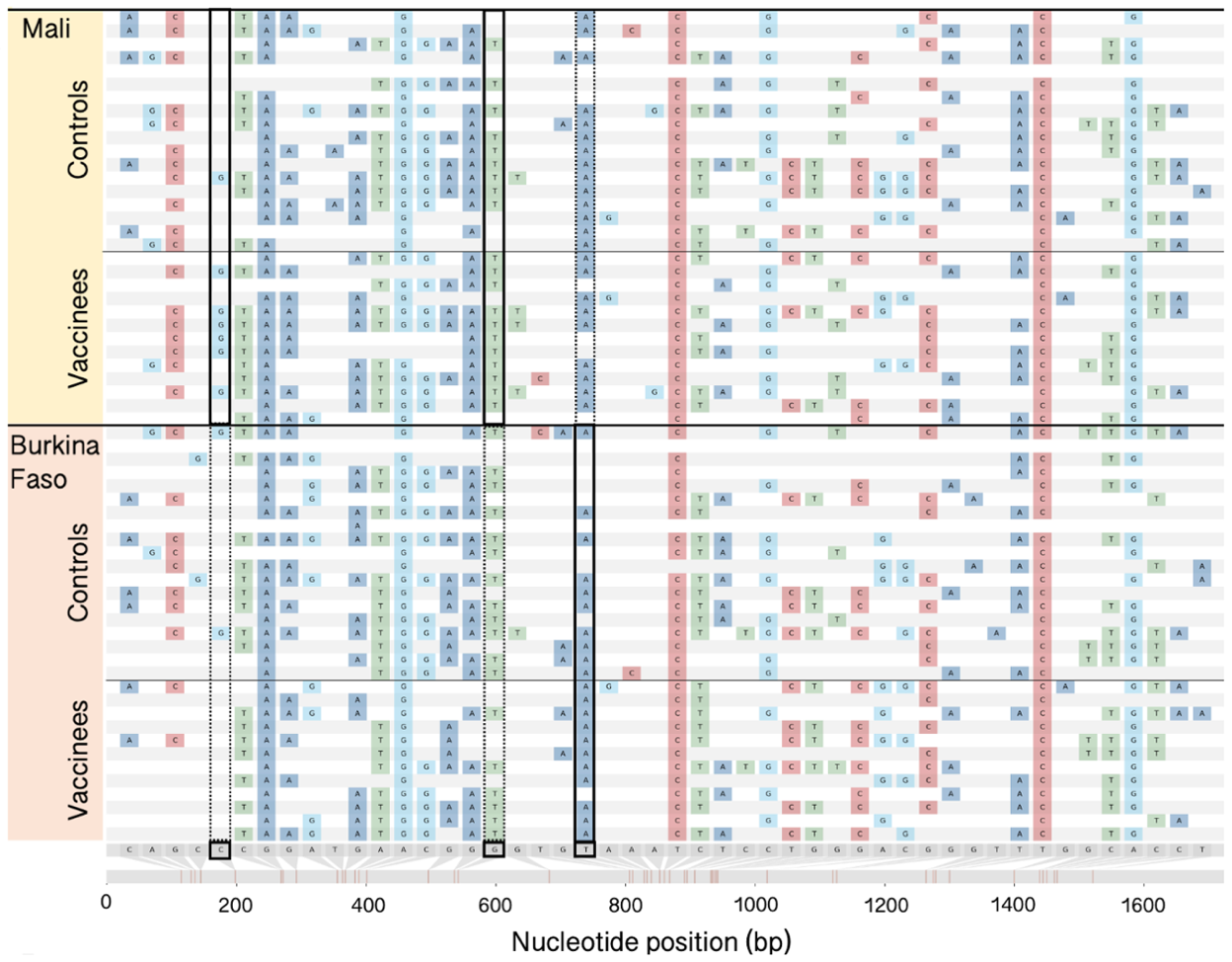

**Figure S6. Genetic variation and distribution of target sites at PF3D7_1335900, the locus encoding PfSSP2|TRAP.** Variable genomic sites in the coding sequence of PF3D7_1335900 among clinical isolates (rows, alternating in white, pink) from vaccinees and controls, from each clinical trial. 3D7 was used as reference sequence and its allelic state at each variable is shown below the alignment (grey). In sequences for each clinical isolate (rows), nucleotides (shown in different colors) denote differences from the 3D7 allelic state; if identical to reference allele, no nucleotide is shown. Target sites identified by SA_WG_ in at least one study are shown (solid box; dotted box in study where site is not a target site). Target sites in the Malian study are not significantly differentiated between vaccinee and control infections in the Burkina Faso trial samples, and vice-versa. Alignment visualization generated with SNIPIT by A. O'Toole. (GitHub repository, 2024).

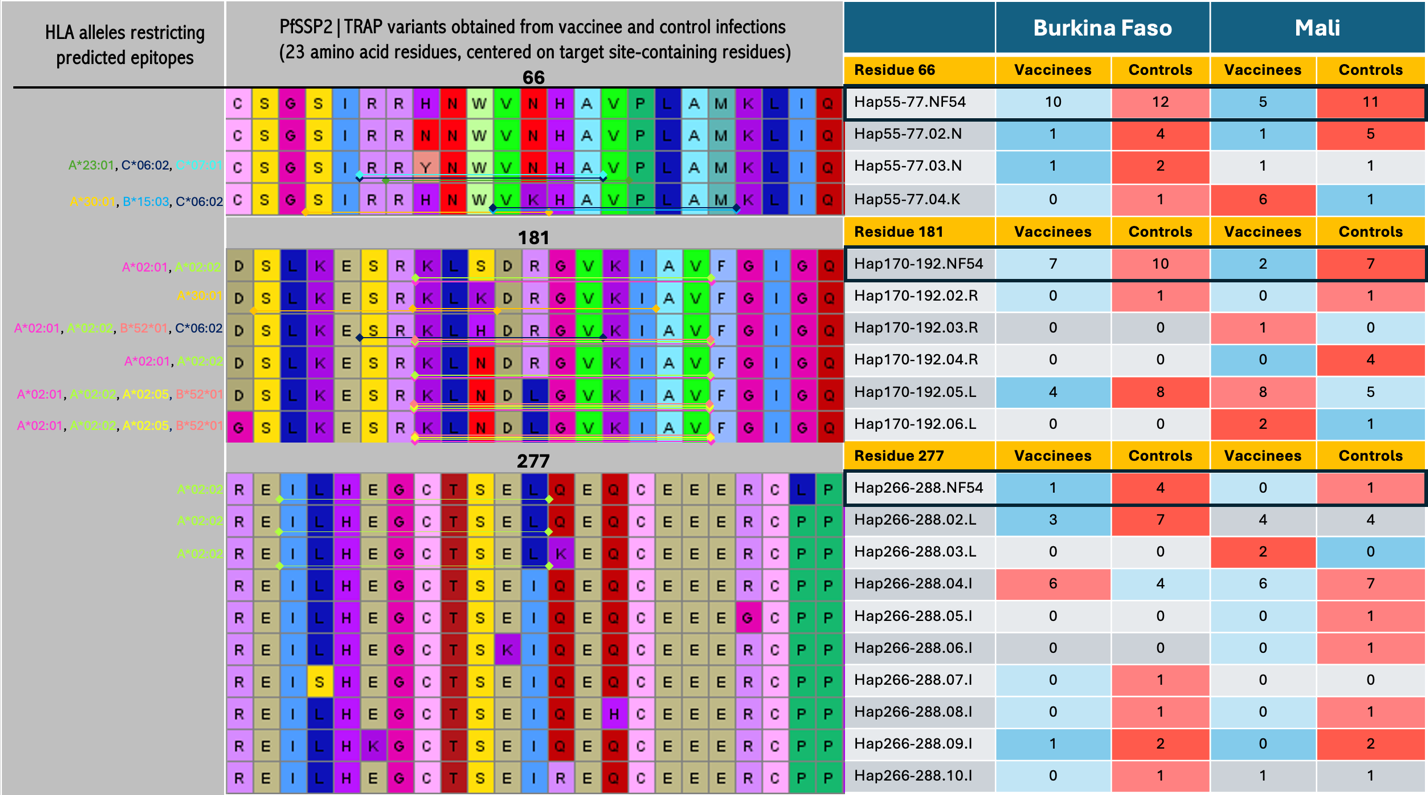

**Figure S7. Distribution of peptide sequences and epitopes surrounding target sites, by study arm, in PF3D7_1335900, the locus encoding PfSSP2|TRAP.** Alignment of TRAP peptide sequences at and surrounding the three residues containing each of the three target sites (central panel). Location of epitope restricted to each HLA allele (left panel) is marked with line across residues forming the CD8 T cell epitope, with color matching between HLA allele and epitope. Frequency of each variant identified in Pf infections from vaccinees or controls is shown (right panel). Vaccine peptide in black frame. Color-coding: same color in both arms when a variant was found in equal number in vaccinees and controls; strong red/blue when frequence was double of more (red) in one arm than the other (blue); light blue/red otherwise.

**Supplemental Tables**

**Table S1. Whole genome sequencing data statistics.** Sample ID and study arm (placebo or vaccine) are shown. DNA sample used for each library was either total DNA or DNA that underwent selective whole genome amplification (sWGA). Sequencing data generated from each genomic library is shown. Total reads generated as well as the number and proportion that mapped to the *P. falciparum* 3D7 genome are shown. The last two columns reflect the proportion of the Pf3D7 genome with at least 1X and 5X read coverage from each library. When two libraries were built for the same sample, data were also merged and processed jointly. Data in green rows was used for downstream analysis. Some samples did not yield sufficient Pf3D7 genome coverage and were not used further (blue rows).

| **Sample ID^1^** | **Study arm** | **DNA sample^2^** | **Total reads sequenced** | **Total reads mapped to**  ***P. falciparum* 3D7** | **Fraction parasite**  **reads ()** | **Genome coverage, >0X (%)** | **Genome coverage, ≥5X (%)** |
| --- | --- | --- | --- | --- | --- | --- | --- |
| **Burkina Faso Sample Set** | | | | | | | |
| IGS-BFA-v0001 | Placebo | Total DNA | 18,514,740 | 11,713,489 | 63.3 | 97.73 | 96.24 |
| IGS-BFA-v0002 | Placebo | Total DNA | 17,464,628 | 8,416,129 | 48.2 | 96.80 | 94.01 |
| IGS-BFA-v0003 | Placebo | Total DNA | 18,056,992 | 10,216,942 | 56.6 | 97.49 | 95.44 |
| IGS-BFA-v0004 | Placebo | Total DNA | 17,916,438 | 13,232,037 | 73.9 | 98.69 | 97.65 |
| IGS-BFA-v0005 | Placebo | Total DNA | 78,169,516 | 16,753,481 | 21.4 | 97.74 | 94.91 |
| IGS-BFA-v0006 | Vaccine | Total DNA | 63,917,666 | 21,376,224 | 33.4 | 96.59 | 92.62 |
| IGS-BFA-v0007 |  | Total DNA | 18,923,480 | 961,110 | 5.1 | 79.40 | 47.68 |
| IGS-BFA-v0007sA |  | sWGA | 29,114,628 | 27,757,039 | 95.3 | 94.75 | 92.67 |
| IGS-BFA-v0007_merged | Placebo | Merged | 48,038,108 | 28,718,149 | 59.8 | 97.87 | 95.52 |
| IGS-BFA-v0008 |  | Total DNA | 52,229,192 | 3,077,790 | 5.9 | 90.17 | 76.00 |
| IGS-BFA-v0008sA |  | sWGA | 26,896,262 | 25,543,863 | 95.0 | 94.82 | 93.34 |
| IGS-BFA-v0008_merged | Placebo | Merged | 79,125,454 | 28,621,653 | 36.2 | 98.02 | 96.77 |
| IGS-BFA-v0009 | Placebo | Total DNA | 88,606,128 | 16,180,330 | 18.3 | 98.24 | 95.73 |
| IGS-BFA-v0010 |  | Total DNA | 26,195,826 | 106,302 | 0.4 | 27.62 | 1.98 |
| IGS-BFA-v0010sA |  | sWGA | 31,044,226 | 29,547,693 | 95.2 | 65.88 | 56.37 |
| IGS-BFA-v0010_merged | Placebo | Merged | 57,240,052 | 29,653,995 | 51.8 | 75.20 | 58.31 |
| IGS-BFA-v0011 |  | Total DNA | 17,935,522 | 1,884,438 | 10.5 | 91.20 | 73.10 |
| IGS-BFA-v0011sA |  | sWGA | 29,692,848 | 28,367,396 | 95.5 | 95.44 | 94.41 |
| IGS-BFA-v0011_merged | Placebo | Merged | 47,628,370 | 30,251,834 | 63.5 | 98.10 | 96.90 |
| IGS-BFA-v0012 | Placebo | Total DNA | 43,103,474 | 21,685,788 | 50.3 | 98.17 | 96.52 |
| IGS-BFA-v0013 |  | Total DNA | 18,094,134 | 46,320 | 0.3 | 1.50 | 0.58 |
| IGS-BFA-v0013sA |  | sWGA | 27,694,200 | 19,280 | 0.1 | 0.56 | 0.21 |
| IGS-BFA-v0013_merged | Vaccine | Merged | 45,788,334 | 65,600 | 0.1 | 1.63 | 0.67 |
| IGS-BFA-v0014 | Placebo | Total DNA | 89,082,708 | 16,947,540 | 19.0 | 98.16 | 96.43 |
| IGS-BFA-v0016 | Vaccine | Total DNA | 17,031,548 | 5,310,264 | 31.2 | 97.08 | 93.63 |
| IGS-BFA-v0017 | Vaccine | Total DNA | 86,675,506 | 15,701,424 | 18.1 | 97.80 | 95.23 |
| IGS-BFA-v0018 | Vaccine | Total DNA | 20,377,570 | 11,028,459 | 54.1 | 97.64 | 96.21 |
| IGS-BFA-v0019 | Vaccine | Total DNA | 25,616,608 | 7,488,257 | 29.2 | 96.68 | 91.30 |
| IGS-BFA-v0021 |  | Total DNA | 16,645,010 | 1,074,464 | 6.5 | 88.68 | 59.81 |
| IGS-BFA-v0021sA |  | sWGA | 26,654,412 | 25,472,579 | 95.6 | 94.60 | 93.36 |
| IGS-BFA-v0021_merged | Placebo | Merged | 43,299,422 | 26,547,043 | 61.3 | 96.98 | 95.33 |
| IGS-BFA-v0022 | Vaccine | Total DNA | 92,051,086 | 20,310,367 | 22.1 | 98.81 | 98.19 |
| IGS-BFA-v0023 | Placebo | Total DNA | 18,566,384 | 8,586,394 | 46.2 | 98.21 | 96.27 |
| IGS-BFA-v0024 |  | Total DNA | 16,732,400 | 470,959 | 2.8 | 64.40 | 21.18 |
| IGS-BFA-v0024sA |  | sWGA | 31,174,744 | 29,711,609 | 95.3 | 94.23 | 92.37 |
| IGS-BFA-v0024_merged | Placebo | Merged | 47,907,144 | 30,182,568 | 63.0 | 95.97 | 93.60 |
| IGS-BFA-v0025 | Placebo | Total DNA | 15,533,572 | 7,513,184 | 48.4 | 97.90 | 95.74 |
| IGS-BFA-v0026 | Placebo | Total DNA | 16,916,132 | 10,896,509 | 64.4 | 97.62 | 96.21 |
| IGS-BFA-v0027 | Placebo | Total DNA | 15,656,672 | 7,073,263 | 45.2 | 97.33 | 95.17 |
| IGS-BFA-v0028 | Vaccine | Total DNA | 22,438,028 | 15,274,336 | 68.1 | 97.66 | 95.90 |
| IGS-BFA-v0029 |  | Total DNA | 59,656,414 | 2,358,915 | 4.0 | 88.16 | 72.44 |
| IGS-BFA-v0029sA |  | sWGA | 26,148,690 | 24,833,085 | 95.0 | 94.61 | 92.76 |
| IGS-BFA-v0029_merged | Vaccine | Merged | 85,805,104 | 27,192,000 | 31.7 | 98.13 | 96.69 |
| IGS-BFA-v0030 |  | Total DNA | 22,657,434 | 1,517,591 | 6.7 | 82.32 | 61.08 |
| IGS-BFA-v0030sA |  | sWGA | 29,283,388 | 27,946,875 | 95.4 | 90.01 | 83.29 |
| IGS-BFA-v0030_merged | Placebo | Merged | 51,940,822 | 29,464,466 | 56.7 | 96.06 | 91.39 |
| IGS-BFA-v0031 |  | Total DNA | 24,460,050 | 2,106,229 | 8.6 | 91.14 | 75.81 |
| IGS-BFA-v0031sA |  | sWGA | 27,240,974 | 25,802,973 | 94.7 | 91.38 | 85.82 |
| IGS-BFA-v0031_merged | Vaccine | Merged | 51,701,024 | 27,909,202 | 54.0 | 97.01 | 94.51 |
| IGS-BFA-v0032 | Vaccine | Total DNA | 17,919,578 | 14,129,837 | 78.9 | 98.88 | 97.79 |
| IGS-BFA-v0033 | Vaccine | Total DNA | 20,744,372 | 17,650,145 | 85.1 | 98.10 | 96.72 |
| IGS-BFA-v0034 | Vaccine | Total DNA | 41,175,804 | 30,890,811 | 75.0 | 98.65 | 97.82 |
| IGS-BFA-v0036 | Placebo | Total DNA | 21,545,606 | 19,759,746 | 91.7 | 98.24 | 97.23 |
| **Mali Sample Set** | | | | | | | |
| IGS-MLI-v0001sA | Placebo | sWGA | 21,874,286 | 20,494,282 | 93.7 | 94.80 | 92.74 |
| IGS-MLI-v0002 |  | Total DNA | 28,260,928 | 1,679,659 | 5.9 | 79.40 | 58.76 |
| IGS-MLI-v0002sA |  | sWGA | 21,211,866 | 20,211,116 | 95.3 | 95.45 | 94.15 |
| IGS-MLI-v0002-merged | Placebo | Merged | 49,472,794 | 21,890,775 | 44.2 | 98.22 | 97.00 |
| IGS-MLI-v0003 |  | Total DNA | 19,044,424 | 172,771 | 0.9 | 31.02 | 5.66 |
| IGS-MLI-v0003sA |  | sWGA | 20,898,778 | 19,920,480 | 95.3 | 94.72 | 93.28 |
| IGS-MLI-v0003-merged | Placebo | Merged | 39,943,202 | 20,093,251 | 50.3 | 96.69 | 94.34 |
| IGS-MLI-v0004 |  | Total DNA | 33,167,608 | 1,642,573 | 5.0 | 80.05 | 58.01 |
| IGS-MLI-v0004sA |  | sWGA | 23,243,940 | 22,231,651 | 95.6 | 95.83 | 94.42 |
| IGS-MLI-v0004-merged | Placebo | Merged | 56,411,548 | 23,874,224 | 42.3 | 98.59 | 97.18 |
| IGS-MLI-v0005 |  | Total DNA | 30,579,442 | 1,295,489 | 4.2 | 75.22 | 51.62 |
| IGS-MLI-v0005sA |  | sWGA | 20,830,396 | 19,957,039 | 95.8 | 95.67 | 94.13 |
| IGS-MLI-v0005-merged | Vaccine | Merged | 51,409,838 | 21,252,528 | 41.3 | 98.46 | 96.83 |
| IGS-MLI-v0006 |  | Total DNA | 49,565,770 | 135,947 | 0.3 | 34.76 | 3.28 |
| IGS-MLI-v0006sA |  | sWGA | 25,299,862 | 24,121,181 | 95.3 | 94.95 | 93.53 |
| IGS-MLI-v0006-merged | Placebo | Merged | 74,865,632 | 24,257,128 | 32.4 | 96.75 | 94.17 |
| IGS-MLI-v0007 |  | Total DNA | 25,812,280 | 180,782 | 0.7 | 37.73 | 5.91 |
| IGS-MLI-v0007sA |  | sWGA | 20,407,556 | 19,473,023 | 95.4 | 94.58 | 92.26 |
| IGS-MLI-v0007-merged | Placebo | Merged | 46,219,836 | 19,653,805 | 42.5 | 96.62 | 93.34 |
| IGS-MLI-v0008sA | Placebo | sWGA | 29,551,982 | 27,979,158 | 94.7 | 94.66 | 93.22 |
| IGS-MLI-v0009 | Vaccine | Total DNA | 151,981,602 | 14,771,415 | 9.7 | 95.58 | 88.79 |
| IGS-MLI-v0010sA | Vaccine | sWGA | 24,095,676 | 12,190,480 | 50.6 | 3.21 | 2.50 |
| IGS-MLI-v0011 | Vaccine | Total DNA | 151,209,760 | 13,446,141 | 8.9 | 95.99 | 91.17 |
| IGS-MLI-v0012sA | Vaccine | sWGA | 24,573,072 | 23,220,131 | 94.5 | 94.92 | 93.19 |
| IGS-MLI-v0013sA | Placebo | sWGA | 23440924 | 22134825 | 94.4 | 94.30 | 91.80 |
| IGS-MLI-v0014 | Placebo | Total DNA | 25,482,448 | 22,625,469 | 88.8 | 98.39 | 97.41 |
| IGS-MLI-v0015 | Vaccine | Total DNA | 88,338,856 | 22,718,544 | 25.7 | 95.53 | 90.65 |
| IGS-MLI-v0017sA | Vaccine | sWGA | 26,240,146 | 24,818,985 | 94.6 | 95.11 | 93.56 |
| IGS-MLI-v0022sA | Placebo | Total DNA | 42,504,564 | 39,651,529 | 93.3 | 89.77 | 83.17 |
| IGS-MLI-v0024 |  | Total DNA | 38,026,220 | 1,222,926 | 3.2 | 63.72 | 40.80 |
| IGS-MLI-v0024sA |  | sWGA | 24,579,978 | 23,525,214 | 95.7 | 95.34 | 94.15 |
| IGS-MLI-v0024-merged | Vaccine | Merged | 62,606,198 | 24,748,140 | 39.5 | 97.64 | 96.55 |
| IGS-MLI-v0027 |  | Total DNA | 52,324,592 | 709,459 | 1.4 | 72.54 | 37.84 |
| IGS-MLI-v0027sA |  | sWGA | 28,078,438 | 26,877,646 | 95.7 | 95.11 | 93.90 |
| IGS-MLI-v0027-merged | Vaccine | Merged | 80,403,030 | 27,587,105 | 34.3 | 97.50 | 95.61 |
| IGS-MLI-v0029 |  | Total DNA | 61,864,116 | 1,161,147 | 1.9 | 73.00 | 48.37 |
| IGS-MLI-v0029sA |  | sWGA | 21,652,902 | 20,764,219 | 95.9 | 95.62 | 94.12 |
| IGS-MLI-v0029-merged | Placebo | Merged | 83,517,018 | 21,925,366 | 26.3 | 97.82 | 96.23 |
| IGS-MLI-v0035 |  | Total DNA | 21,358,200 | 12,149 | 0.1 | 2.20 | 0.09 |
| IGS-MLI-v0035sA |  | sWGA | 19,874,652 | 18,980,192 | 95.5 | 57.01 | 45.48 |
| IGS-MLI-v0035-merged | Placebo | Merged | 41,232,852 | 18,992,341 | 46.1 | 58.02 | 45.63 |
| IGS-MLI-v0041 | Placebo | Total DNA | 71,423,646 | 17,104,835 | 23.9 | 96.81 | 91.47 |
| IGS-MLI-v0042sA | Vaccine | sWGA | 36,943,364 | 35,033,617 | 94.8 | 95.04 | 93.94 |
| IGS-MLI-v0043sA | Placebo | sWGA | 21,977,460 | 20,741,020 | 94.4 | 94.48 | 92.90 |
| IGS-MLI-v0044 | Vaccine | Total DNA | 132,739,214 | 20,449,806 | 15.4 | 96.68 | 91.66 |
| IGS-MLI-v0045sA | Placebo | sWGA | 20,768,774 | 19,637,604 | 94.6 | 94.99 | 93.29 |
| IGS-MLI-v0048 |  | Total DNA | 34,778,268 | 895,481 | 2.6 | 73.81 | 43.01 |
| IGS-MLI-v0048sA |  | sWGA | 20,873,630 | 19,963,680 | 95.6 | 95.58 | 94.22 |
| IGS-MLI-v0048-merged | Vaccine | Merged | 55,651,898 | 20,859,161 | 37.5 | 98.19 | 96.50 |
| IGS-MLI-v0050 |  | Total DNA | 39,936,356 | 599,531 | 1.5 | 65.36 | 30.98 |
| IGS-MLI-v0050sA |  | sWGA | 17,083,300 | 16,361,839 | 95.8 | 94.75 | 93.22 |
| IGS-MLI-v0050-merged | Placebo | Merged | 57,019,656 | 16,961,370 | 29.7 | 97.07 | 95.07 |
| IGS-MLI-v0057 |  | Total DNA | 43,043,824 | 25,870 | 0.1 | 8.99 | 0.04 |
| IGS-MLI-v0057sA |  | sWGA | 55,401,070 | 28,063,165 | 50.7 | 94.88 | 92.25 |
| IGS-MLI-v0057-merged | Vaccine | Merged | 98,444,894 | 28,089,035 | 28.5 | 95.25 | 92.32 |
| IGS-MLI-v0058 |  | Total DNA | 29,872,988 | 13,588 | 0.0 | 4.90 | 0.01 |
| IGS-MLI-v0058sA |  | sWGA | 36,739,900 | 23,465,176 | 63.9 | 94.65 | 93.05 |
| IGS-MLI-v0058-merged | Vaccine | Merged | 66,612,888 | 23,478,764 | 35.2 | 94.86 | 93.07 |
| IGS-MLI-v0060sA | Placebo | sWGA | 57,193,198 | 43,912,950 | 76.8 | 87.53 | 77.38 |
| IGS-MLI-v0061 |  | Total DNA | 23,149,658 | 11,437 | 0.0 | 4.20 | 0.001 |
| IGS-MLI-v0061sA |  | sWGA | 55,123,280 | 39,561,165 | 71.8 | 74.00 | 55.81 |
| IGS-MLI-v0061-merged | Vaccine | Merged | 78,272,938 | 39,572,602 | 50.6 | 75.06 | 56.06 |
| IGS-MLI-v0062 |  | Total DNA | 25,822,536 | 38,194 | 0.1 | 13.58 | 0.24 |
| IGS-MLI-v0062sA |  | sWGA | 24,522,746 | 19,502,903 | 79.5 | 95.38 | 93.56 |
| IGS-MLI-v0062-merged | Placebo | Merged | 50,345,282 | 19,541,097 | 38.8 | 95.90 | 93.69 |
| IGS-MLI-v0064 |  | Total DNA | 32,138,602 | 85,359 | 0.3 | 26.15 | 0.76 |
| IGS-MLI-v0064sA |  | sWGA | 22,284,810 | 110 | 0.0 | 0.05 | NA |
| IGS-MLI-v0064-merged | Vaccine | Merged | 54,423,412 | 85,469 | 0.2 | 26.19 | 0.76 |
| IGS-MLI-v0065 |  | Total DNA | 23,298,232 | 15,746 | 0.1 | 5.76 | 0.02 |
| IGS-MLI-v0065sA |  | sWGA | 17,262,200 | 1,064,123 | 6.2 | 79.06 | 48.28 |
| IGS-MLI-v0065-merged | Placebo | Merged | 40,560,432 | 1,079,869 | 2.7 | 80.18 | 48.96 |
| IGS-MLI-v0067 |  | Total DNA | 52,431,816 | 890,152 | 1.7 | 64.58 | 24.87 |
| IGS-MLI-v0067sA |  | sWGA | 24,103,766 | 23,140,621 | 96.0 | 95.08 | 92.85 |
| IGS-MLI-v0067-merged | Placebo | Merged | 76,535,582 | 24,030,773 | 31.4 | 98.14 | 96.49 |
| IGS-MLI-v0068 |  | Total DNA | 32,347,604 | 15,346 | 0.0 | 5.45 | 0.02 |
| IGS-MLI-v0068sA |  | sWGA | 76,305,704 | 21,617,380 | 28.3 | 65.76 | 47.44 |
| IGS-MLI-v0068-merged | Vaccine | Merged | 108,653,308 | 21,632,726 | 19.9 | 67.55 | 47.79 |

**Table S2. Number and distribution of polymorphic/variable sites (VS) in samples from each study.**

|  | Burkina Faso  PfSPZ Vaccine study | | Mali  PfSPZ CVac study | | Union set between studies |
| --- | --- | --- | --- | --- | --- |
|  | Nuclear genome | Nuclear genome, Core regions only | Nuclear genome | Nuclear genome, Core regions only | Nuclear genome |
| VS, Total | 31,122 | 28,274 | 31,659 | 30,193 | 39,458 |
| VS in protein-coding loci | 20,547 | 18,702 | 19,776 | 18,977 | 24,965 |
| NSYN VS | 12,678 | 11,701 | 12,136 | 11,686 | 15,198 |
| Genes with VS | 3,988 | 3,797 | 4,043 | 3,929 | 4,324 |
| Genes w NSYN VS | 3,064 | 2,891 | 3,033 | 2,939 | 3,326 |
| Single copy genes with NSYN VS | 2,603 | 2,600 | 2,649 | 2,646 | 2,848 |

**Table S3. Significantly differentiated genomic sites in Burkina Faso PfSPZ vaccine trial.** The genomic coordinates, Wier and Cockerham’s *F*_ST_ value and gene descriptions for sites with *p-value* (permutation method; k=5000) < 0.05 is shown. The *F*_ST_ value is calculated for each site between Placebo (n=19) and Vaccinee (n=12) samples, using vcftools. Additional columns indicate whether the NF54 allele is depleted among vaccinee samples and whether a SNP is present in known pre-erythrocytic vaccine antigens. The rows are highlighted to show target sites that fall in target loci common to both studies (blue; n=18) and specific target sites that appear in both studies (orange; n=2). See Excel spreadsheet Table S3.

**Table S4. Significantly differentiated genomic sites in Mali PfSPZ CVac vaccine trial.** The genomic coordinates, Wier and Cockerham’s *F*_ST_ value and gene descriptions for sites with *p-value* (permutation method; k=5000) < 0.05 is shown. The *F*_ST_ value is calculated for each site between Placebo (n=18) and Vaccinees (n=13) cohort, using vcftools. Additional details as in Supplemental Table S3. See Excel spreadsheet Table S4.

**Table S5. NF54 allele depletion by study arm, for significantly differentiated non-synonymous (NSYN) sites**. Statistical significance was determined using a chi-square test.

| Study | Vaccine allele depleted in controls  n (%) | Vaccine allele depleted in vaccinees  n (%) | Significantly differentiated NSYN sites  n | *p*-value |
| --- | --- | --- | --- | --- |
| Burkina Faso | 60 (22.9) | 202 (77.1) | 262 | **< 0.001** |
| Mali | 69 (38.5) | 110 (61.5) | 179 | **< 0.001** |

**Table S6. Distribution of target sites relative to CD8+ T cell epitopes, in the twelve single-copy target loci common to both studies.** The location of strong-binding epitopes in the twelve target loci of interest was determine for each of the 22 most prevalent HLA alleles known in Burkina Faso and Mali. The distribution of non-synonymous target sites among the reunion of all strong-binding CD8+ T cell epitopes (CD8+ immuno-peptidome) *versus* outside of this immune-peptidome was determined. Statistical significance for cumulative distribution was assessed with a chi-square test.

|  | | Data partition | | |  | |  |
| --- | --- | --- | --- | --- | --- | --- | --- |
|  | | Sites in CD8+ T cell immuno-peptidome | Sites not in immuno-peptidome | | Total | | *p*-value |
| Observed NSYN target sites | | 22 | 5 | | 27 | | 0.74 |
| Expected NSYN target sites | | 20 | 7 | | 27 | |  |
| Cumulative number of non-synonymous sites in 12 target loci (bp) | | 47111 | 15180 | | 62291 | |  |
| Distribution of nonsynonymous (NSYN) target sites per locus | | | | | | | |
| **Gene ID** | **Gene product** | | | Sites in CD8+ T cell immuno-peptidome | | Sites not in immuno-peptidome | |
| PF3D7_0113800 | DBL-containing protein | | | 3 | | 0 | |
| PF3D7_0421700 | Conserved Plasmodium protein | | | 2 | | 0 | |
| PF3D7_0518700 | mRNA-binding protein PUF1 | | | 2 | | 0 | |
| PF3D7_0609600 | Conserved Plasmodium protein | | | 2 | | 0 | |
| PF3D7_0703900 | Conserved Plasmodium membrane protein | | | 3 | | 0 | |
| PF3D7_0711200 | Conserved Plasmodium protein | | | 1 | | 1 | |
| PF3D7_0713600 | Ribosomal protein S5, mitochondrial, putative | | | 1 | | 1 | |
| PF3D7_0808100 | AP-3 Complex subunit Delta | | | 2 | | 0 | |
| PF3D7_0826000 | Conserved Plasmodium protein | | | 2 | | 0 | |
| PF3D7_1324300 | Conserved Plasmodium membrane protein | | | 1 | | 1 | |
| PF3D7_1335900 | Thrombospondin-related anonymous protein, TRAP | | | 2 | | 1 | |
| PF3D7_1361800 | Glideosome-associated connector, GAC | | | 1 | | 1 | |

**Table S7. Samples used for joint SNP calling.** List of 1,377 samples used in this study, with available short-read data (Illumina), for joint SNP calling, including those sequenced as part of this study and those publicly available data.
